# A Case of Excessive, Opportunistic Price Increase of a Generic Oncology Drug

**DOI:** 10.1101/2025.07.05.25330918

**Authors:** Parthay H Patel, Bhaumik B Patel, Umesh R Desai

## Abstract

**Background:** Off-patent, generic drugs are expected to be widely accessible owing competitive pricing and minimal R&D costs. Sudden, excessive, unexplainable, and sustained price increases detrimentally impact the health and welfare of patients and their families. Overlooking such cases encourages repeat behavior that may be legislatively challenging to overcome, thereby impeding the principle of democratization of medicines propagated by the expiration of patent protection.

**Findings:** We analyzed Medicare Part B quarterly payment data over 2020 to 2025 and observed a sudden, steep and sustained increase of over 5000% in the price of floxuridine injection, with the most noticeable spike between October 2022 and January 2023. Comparable increases in magnitude were also corroborated in state Medicaid programs. Plausible explanations, including raw material shortage, regulatory changes, formulation changes, and change in patent status, did not support the steep price hike. Market exit of manufacturer(s) to the point of exclusivity appears to be the only reasonable explanation for the price hike.

**Conclusions:** We conclude that although legally permissible, the opportunistic excessive pricing of floxuridine originates from market consolidation and supplier exclusivity. This case reflects a broader concern that generic drugs may be subject to morally questionable pricing. As floxuridine becomes more widely available through ongoing clinical trials, concerns about its affordability and financial toxicity will be difficult to ignore. Floxuridine serves as a cautionary tale of how niche-market generics can evade public scrutiny while imposing unnecessary costs on our healthcare system.

---

The pricing of generic pharmaceutical drugs involves multiple factors including a) exclusivity, or lack of competition, b) geopolitical dynamics of raw material and manufacturing costs, and c) major regulatory changes related to pharmaceutical products, especially active pharmaceutical ingredients. Despite much research, drug pricing and price variation has been challenging to interpret and predict.^1^ One reason for this is that the pricing and reimbursement of drugs operates under a non-transparent system, which hampers assessing the true value of medications, while also impeding fair and equitable access. In fact, when daraprim price was increased by more than 5000%, it was deemed exploitative and morally wrong, especially for those suffering from toxoplasmosis, a rare condition affecting a few thousand patients.^2^ Additionally, the financial barrier dramatically decreased drug access and increased treatment delays and interruptions, especially in HIV/AIDS patients.^3^

We report the case of pricing of another generic drug that appears to mimic the case of daraprim. Floxuridine was first approved by the US FDA in 1970 and is used to treat patients of colorectal cancer.^4^ Although the incidence of colorectal cancer in the US is huge, floxuridine usage is much less (<$1 billion/yr), which disfavors development of a typical competitive market. Alternatively, the smaller, but not small, market combined with supplier exclusivity appears to have induced exploitive pricing, which has gone completely unnoticed.

Floxuridine Injection (HCPCS Code: J9200) is an antimetabolite cytotoxic agent, which is used in the palliative treatment of gastrointestinal adenocarcinoma that has metastasized to th liver.^5^ To evaluate the recent pricing trends of floxuridine, Medicare Part B quarterly payment limit data were obtained from Average Sales Price (ASP) files and analyzed to calculate the percentage changes in price from April 2020 through April 2025.^6^ An extreme 5059% increase in price was noticed for this generic drug over this time period (**Figure 1A**). More strikingly, a steep increase took place within a few months (October 2022 and January 2023) with the payment limit increasing from $90 to $2890 (**Table S1**). This finding is also replicated in Medicaid programs, for example, Montana’s Physician Fee Schedule, which notes an increase from $97 to $3551 between July 2022 and July 2023 (**Table S2**).^7,8^

**Figure 1.**
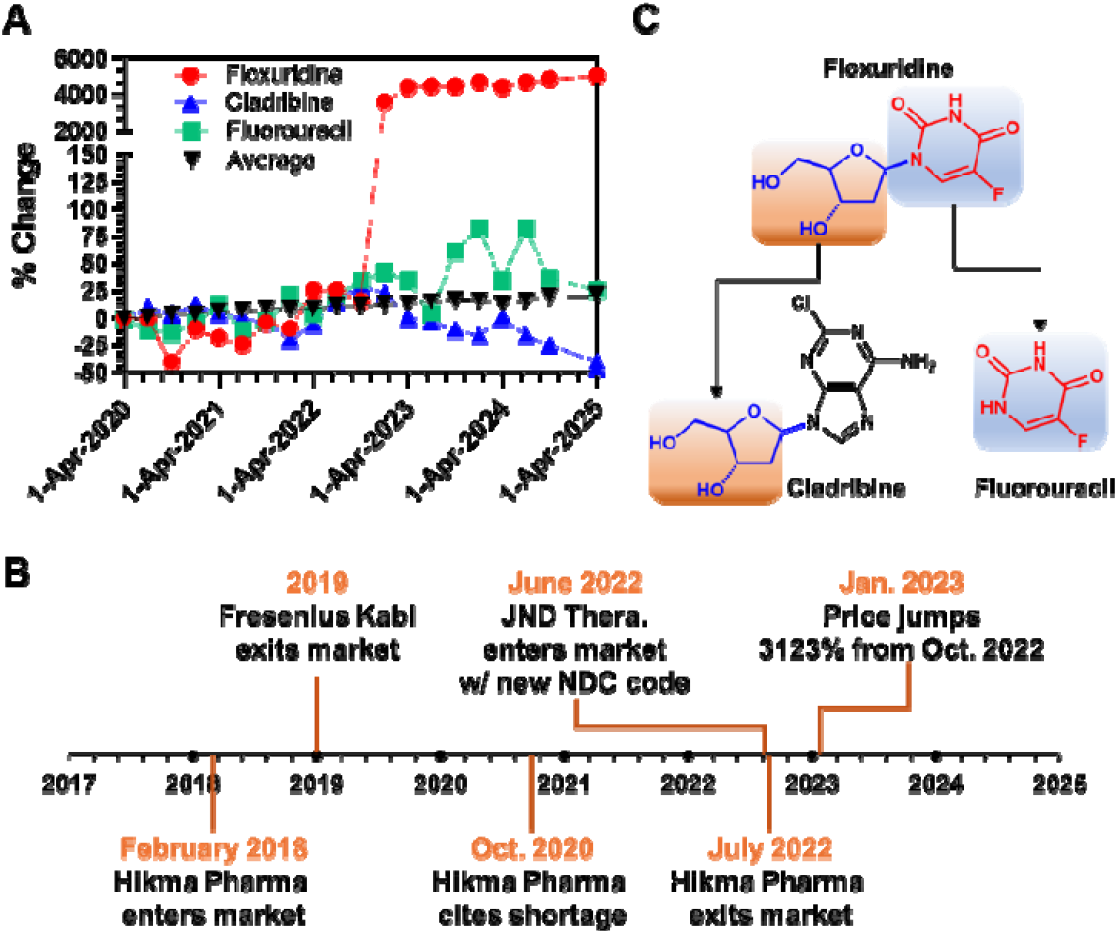
Floxuridine injection price change over 2020–2025. A) Cumulative average sales price (ASP) changes for drugs reimbursed by Medicare Part B from April 2020 to April 2025. Prices were calculated by averaging the ASP for each drug per quarter and then computing the cumulative growth. Only drugs with reported ASP data for all five years were included. Cumulative ASP changes for floxuridine (red), fluorouracil (green), and cladribine (blue) are compared with the overall average of all Medicare Part B drugs (black). Raw data were sourced from Centers for Medicare and Medicaid Services (CMS) quarterly ASP payment limits on 05/22/2025. B) Timeline of floxuridine market events summarizing manufacturer exit/entry, raw material shortage, and price increase. Some dates are approximations due to limited publicly available information. NDC = National Drug Code. C) Structures of three related drugs, floxuridine, cladribine, and fluorouracil. See text for details.

To assess whether this dramatic increase can be rationalized, we studied several plausible factors associated with drug pricing and price fluctuations. One explanation could be a severe shortage of raw materials. This was suggested by the American Society of Health-System Pharmacists in October 2020 owing to drug discontinuation by one manufacturer and a raw material shortage claim by another.^9^ However, this does not adequately explain the price increase of floxuridine in late 2022/early 2023. The price escalation occurred more than two years later. Further, a review of the exit/entrance timeline of manufacturers shows that the steep price hike came about after the only manufacturer’s market exit in 2022, when a third manufacturer entered the market (**Figure 1B**).^10^

We also reasoned that comparing pricing of related drugs during this time period may offer additional insight. Floxuridine is a prodrug, which is metabolized in the body to fluorouracil, the active ingredient. In terms of manufacturing, floxuridine is synthesized by coupling two raw materials including fluorouracil and the deoxyribose sugar (**Figure 1C**). Interestingly, minimal price increase is observed for fluorouracil, the first raw material, over the past 5 years (**Figure 1A**). To assess whether the second raw material could be culprit behind the steep price hike, we studied cladribine, a clinically used antimetabolite and containing the same deoxyribose sugar (**Figure 1C**). No price escalations are observed for cladribine (**Figure 1A**). Thus, neither raw materials used in the manufacture of floxuridine saw steep price hikes, especially during the late 2022/early 2023 time period.

To assess how unusual the floxuridine price increase was, we calculated the change in ASP for all drugs of Medicare Part B over the last 5 years (**Table S3**). The next highest price increase was observed for prednisolone, which presented a substantial, but comparatively modest, 393% increase. In contrast, the average increase in price of Medicare Part B drugs was a meager 21.8% (**Figure 1A**), which compares favorably with the cumulative inflation of ~25% during that time reported by the US Bureau of Labor Statistics.^11^

Another plausible explanation for the dramatic price increase could be formulation and/or regulatory changes introduced by the Food and Drug Administration (FDA) or Centers for Medicare and Medicaid Services (CMS). Over the period of observation, there were no significant changes in regulations related to floxuridine. Likewise, as an off-patent generic drug, floxuridine injection has not seen any change in formulation over the years.^12,13^ On the contrary, the Inflation Reduction Act enacted in 2022 was aimed to reduce drug prices through Medicare price negotiation.^14^

The primary explanation for the steep price hike appears to be market consolidation aided exclusivity (**Figure 1B**). In 2019, Fresenius Kabi stopped producing floxuridine, leaving Hikma Pharmaceutical, who entered in February 2018, as the only producer of the drug.^9,15^ Later, Hikma cited the raw material shortage and exited the market.^9,10^ Cerona Therapeutics, which would later rebrand as JND Pharmaceuticals, then temporarily took upon the distribution of the drug from Hikma.^9^ On June 9, 2022, a new label code for floxuridine injection appeared in FDA’s NDC registry with JND Pharmaceuticals becoming the sole supplier of the drug (**Figure 1B**).^16^ Soon thereafter, massive price increase came about. This suggests exclusivity, or lack of competition (**Table S4**), as the primary driver for this legal, but morally inappropriate, pricing.

At present, floxuridine injection is used in a relatively limited number of centers that offer hepatic intra-arterial pump treatment and usually in a clinical trial for a specific indication of treating unresectable liver metastasis in patients with colorectal and other gastrointestinal cancer. Yet, its pricing is an important consideration now as it may become the new standard in the future based on the outcomes of several ongoing clinical trials.^17^ In fact, current exclusivity of use may have allowed the drug companies to slide this exuberant price increase past the general public’s scrutiny.

Overlooking this excessive and opportunistic price increase should not be an option. Floxuridine may be widely prescribed in the future. More than 250,000 new patients worldwide are likely to become eligible for the treatment assuming success of ongoing clinical trials.^18,19^ As of April 2025, Medicare Part B reimburses a payment limit of $3,988 for floxuridine with typical patients covering a coinsurance of 20%.^6^ Although the total number of patients cannot be accurately estimated until ongoing studies mature, targeting the pricing of niche specialty medications, such as floxuridine, should be a matter of importance and concern for patients, physicians, insurance companies, regulators, and policy makers. It is important to reduce financial toxicity for patients. It is also important to enable access of key drugs to all patients. The fact that generic drugs can also suffer from monopoly-based pricing, which is theoretically important for innovation, should sound an alarm to people and policy makers. It defeats the purpose of the principle of expiration of patent protection. It is imperative that this case of floxuridine and the insights gained from the analysis of plausible explanations behind the excessive and opportunistic pricing be brought out by the stewards of the practitioners of medicine, especially physicians and pharmacists.

## Data Availability

A spreadsheet listing the raw data on drug costs collected from the Centers for Medicare and Medicaid Services (CMS) will be available upon request from the corresponding author.

https://www.cms.gov/medicare/payment/part-b-drugs/asp-pricing-files

https://medicaidprovider.mt.gov/

## Author Contributions

PHP collected and analyzed the raw data, prepared and edited manuscript drafts; BBP and URD interpreted the data and edited manuscript.

