## Supplemental Tables for "A Case of Excessive, Opportunistic Price Increase of a Generic Oncology Drug"

**Table 1. Floxuridine Injection (HCPCS Code: J9200) Price Change Over 2020–2025.*^a^***

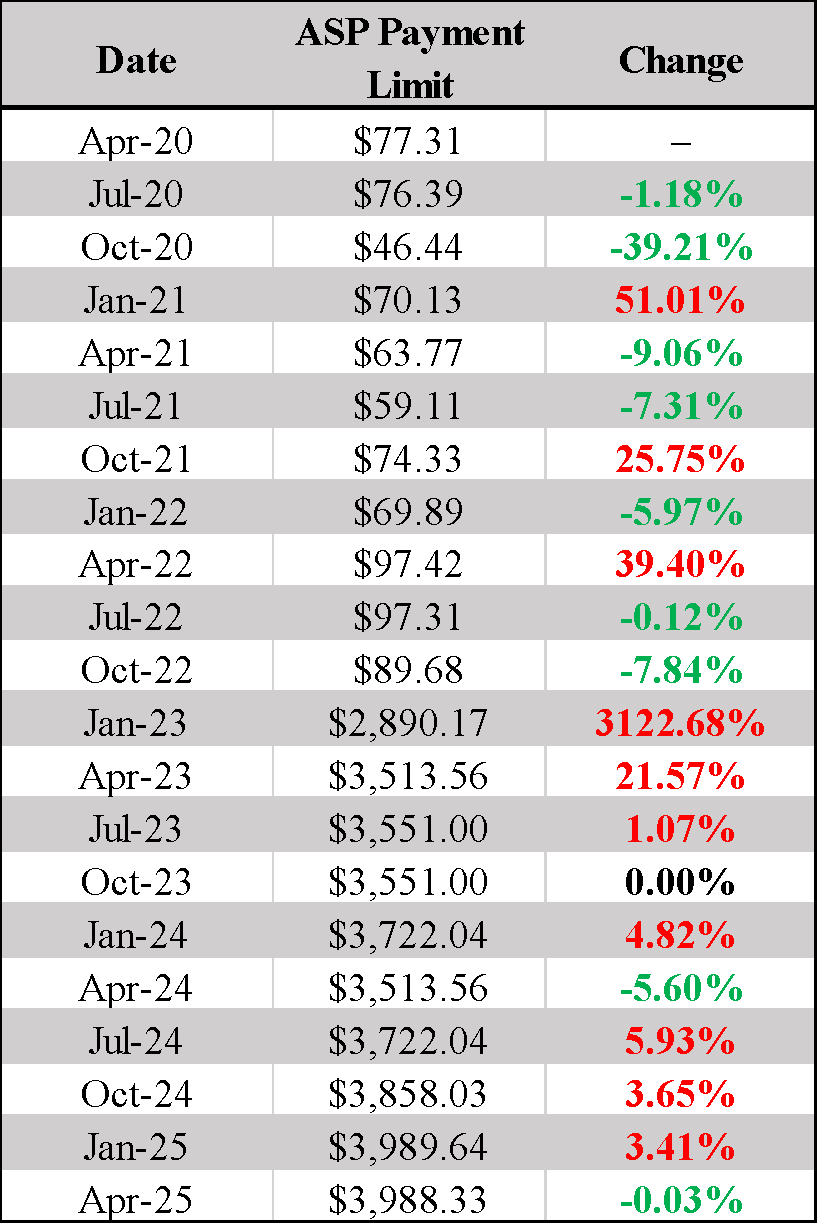

*^a^ Data were sourced from Centers for Medicare and Medicaid Services (CMS) quarterly ASP payment limits.*

*Abbreviations: ASP, average sales price; HCPCS, Healthcare Common Procedure Coding System.*

**Table S2. Floxuridine Injection Price Change at Medicaid Over 2020–2025.*^a^***

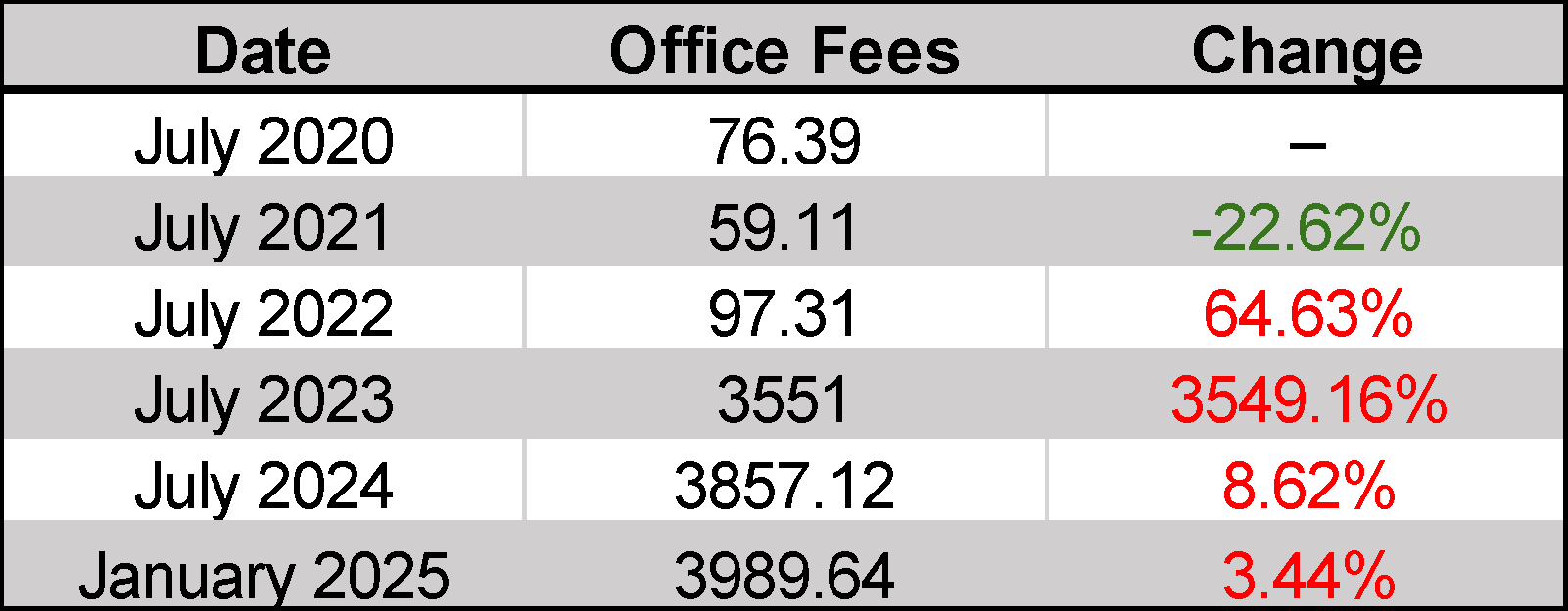

*^a^ Data were sourced from Montana Department of Public Health and Human Services’ Medicaid Physician Fee Schedules for July 2020 – 2024 and January 2025.*

**Table S3. Top 5 Price Hikes in Medicare Part B Drugs (Five-Year Trend).*^a^***

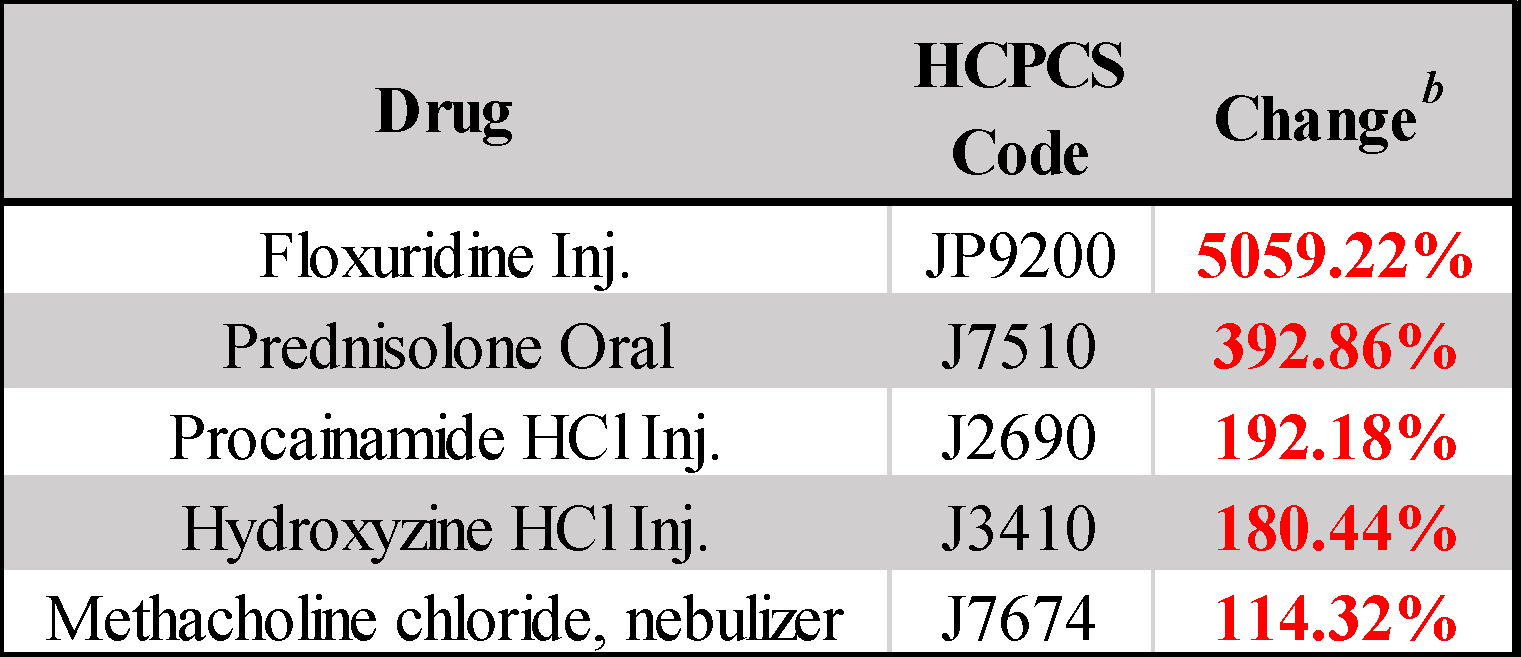

*^a^ Data were sourced from Centers for Medicare and Medicaid Services (CMS) quarterly ASP payment limits.*

*^b^ Price change reflects the five-year cumulative change in Medicare Part B ASP payment limit.*

*Abbreviations: ASP, average sales price; HCPCS, Healthcare Common Procedure Coding System.*

**Table S4. Summary of potential factors behind floxuridine price spike.**

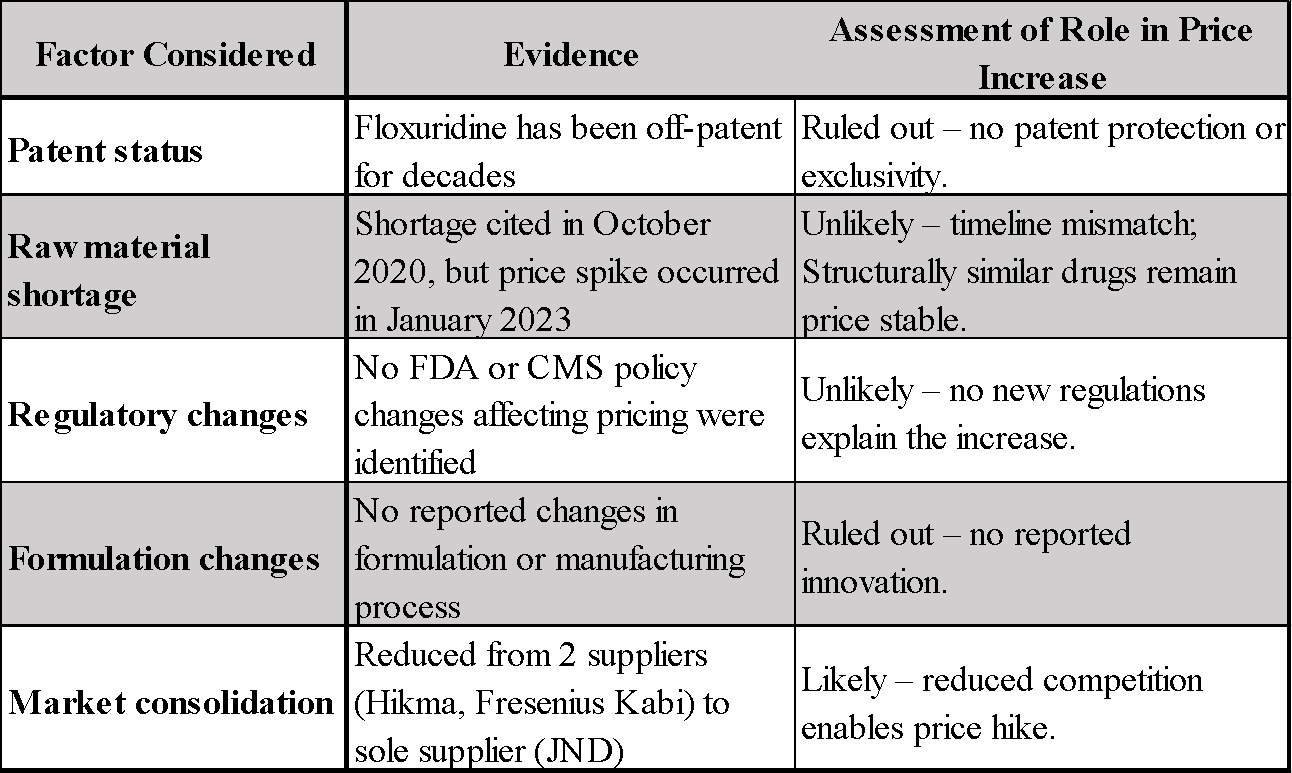

*Abbreviations: FDA, Food and Drug Administration; CMS, Centers for Medicare and Medicaid Services.*
